# Generating intermediate slices with U-nets in craniofacial CT images

**DOI:** 10.1101/2024.05.08.24307089

**Authors:** Soh Nishimoto, Kenichiro Kawai, Koyo Nakajima, Hisako Ishise, Masao Kakibuchi

**Affiliations:** Department of Plastic Surgery, Hyogo Medical University 1-1 Mukogawa-cho, Nishinomiya, Hyogo 663-8131, Japan

**Keywords:** Computer tomography, Facial bone fracture, U-net, Interpolation, Intermediate slice

## Abstract

**Aim:** The Computer Tomography (CT) imaging equipment varies across facilities, leading to inconsistent image conditions. This poses challenges for deep learning analysis using collected CT images. To standardize the shape of the matrix, the creation of intermediate slice images with the same width is necessary. This study aimed to generate inter-slice images from two existing CT images.

**Materials and Methods:** The study utilized CT images from the Japanese Facial Bone Fracture CT Collection Project. The pixel values were converted to Hounsfield numbers and normalized. Three re-slice systems utilizing U-nets were developed: 1/3, 1/4, and 1/5. The datasets were divided into training and validation sets, and data augmentation techniques were applied. The U-net models were trained for 200 epochs. Validation was conducted using validation datasets. The generated images were compared to the corresponding original images using peak signal-to-noise ratio (PSNR), structural similarity (SSIM), and mean squared error (MSE) calculations. Results: Statistical analysis revealed significant differences between linear interpolation and U-net prediction in all indexes.

**Conclusion:** The developed re-slice systems with U-net models showed practical value for making intermediate slice images from the existing images in the craniofacial area.

## Introduction

X-ray CT (Computer Tomography) is an indispensable modality in diagnosing and treating patients, nowadays. Usually, the images are stored in DICOM (Digital Imaging and Communications in Medicine) format. To reduce data size for saving, exclusive drawing out images with thick intervals from the original ones is often done. The other slices with detailed information are discarded. Rebuilding a 3D image from these sparsely selected slices would result in a coarse one.

The CT imaging equipment differs from facility to facility, and the imaging conditions are not standardized. When CT images are collected from multiple facilities, the variety of image conditions can cause problems. In order to perform 3D deep learning from the collected CT images, it is necessary to regularize the shape of the matrix. Though, it is not easy to do so, because the slice width is not the same for each patient. To create sets of images with the same slice width, the creation of the intermediate slice images from the existing images is necessary.

In this study, the generation of the inter-slice images from two existing CT images (re-slicing) was attempted.

## Materials and Methods

All procedures were done on a desk-top personal computer with a GPU: GeForce RTX3090 24.0GB ((nVIDIA, Santa Clara, CA, USA), Windows 10 Pro (Microsoft Corporations, Redmond, WA, USA). Python 3.8(1)(Python Software Foundation, DE USA): a programming language, was used under Anaconda 15 (2)(FedoraProject. http://fedoraproject.org/wiki/Anaconda#Anaconda_Team_Emeritus) as an installing system, and Spyder 4.1.4(2) as an integrated development environment. Keras 3(3)(https://keras.io/): the deep learning library, written in Python was run on TensorFlow 2.5 (Google, Mountain View, CA, USA). GPU computation was employed through CUDA 10.0(4)(nVIDIA). A Python library: pydicom (https://pydicom.github.io) was used to deal with DICOM files.

### Data

CT images (280 cases, 57931 images in total) accumulated in the Japanese Facial Bone Fracture CT Collection Project (Umin000039624) were used. The project has been approved by The Ethics Review Board of Hyogo Medical University (No.3326). The images were taken for the purpose of diagnosing fractures in craniofacial areas. Patients’ written consents for the study use were obtained. They were processed so that the personal information was not identified and stored in DICOM files. Imaging conditions varied.

The images were read with pydicom and pixel values were converted to Hounsfield numbers with apply_modality_lut module. The values below - 1024 and above +1024 were truncated. Then 1024 was added, divided by 2048, and normalized into 0.0 to 1.0. Most of the accumulated images were in axial view, but some were not. They were eliminated by checking the DICOM tags. The localizer images were also eliminated.

### Slice level classifier

A deep learning system to classify what level the CT image had been sliced(5) was previously established. Briefly, the CT images were visually classified into 6 regions: blank, head, orbital, maxilla, mandible, and neck. Randomly, 75% of each site was selected for training and 25% for validation. A ResNet-50(6) system to classify 6 classes was made and trained with data-augmented (zooming, rotation, shifting) training data (Figure 1). The accuracy for training was 0.9909 and for validation was 0.9873.

**Figure 1.**
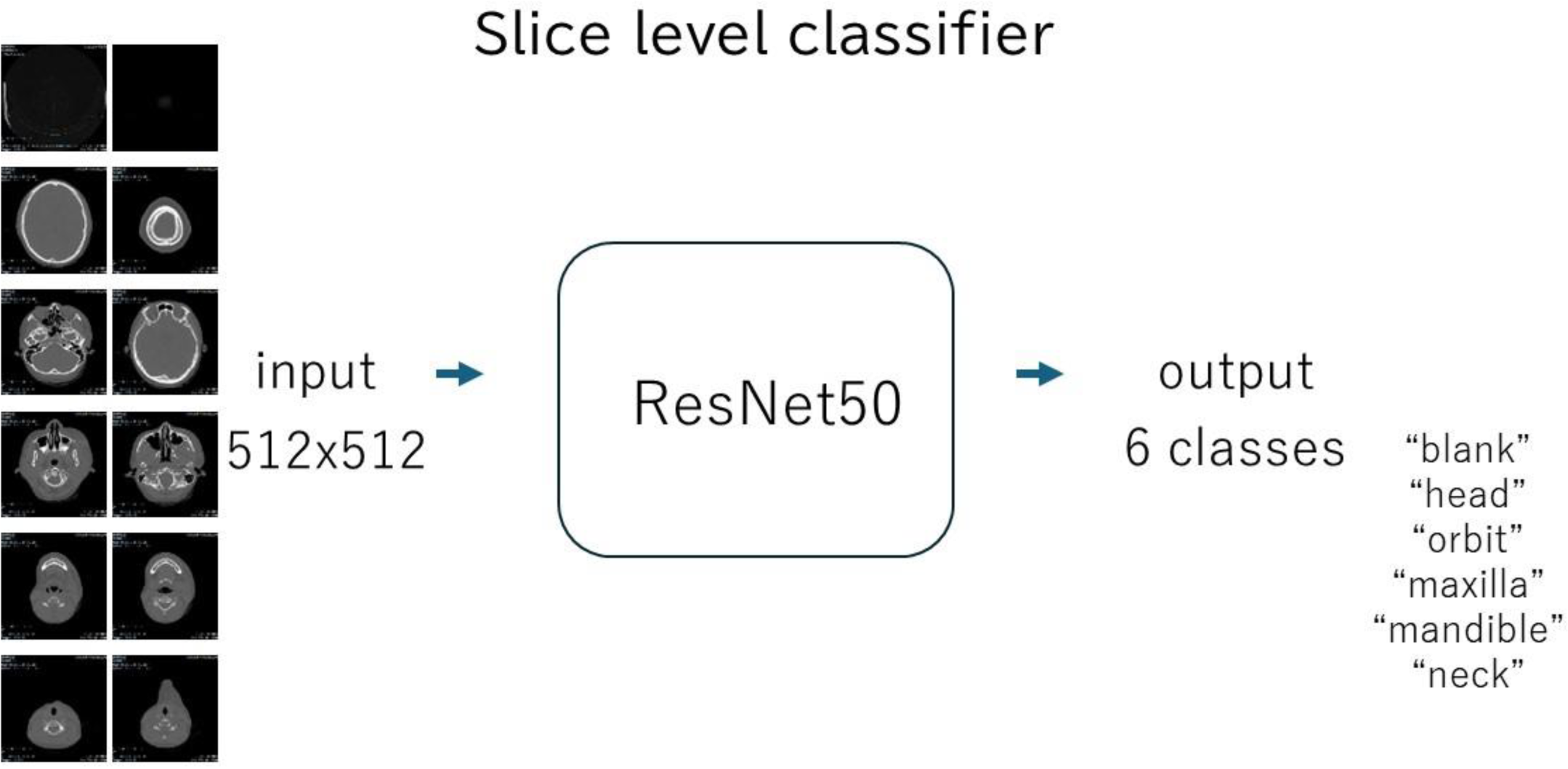
Diagram of the slice level classifier. The trained network was used to search blank images.

A series of CT images for each patient was processed with the slice level classifier, mentioned above. The images classified “blank” were eliminated. The images were aligned in the order of image position. A total of 20457 horizontal sections for 102 cases with no missing data were used in this study. The variation of slice width in the series of CT images, used in this study, is shown in Table1.

### Training U-nets

#### (1) 1/3 re-slice system (Figure 2, 3)

One image and the 4th image (skipping 2 images) were paired. They were stacked (512x512x2) and collected as input. The 2nd and 3rd images stacked (512x512x2) were collected as output. Then, one image and the 7th image counted from it were stacked. That was collected as input.

**Figure 2.**
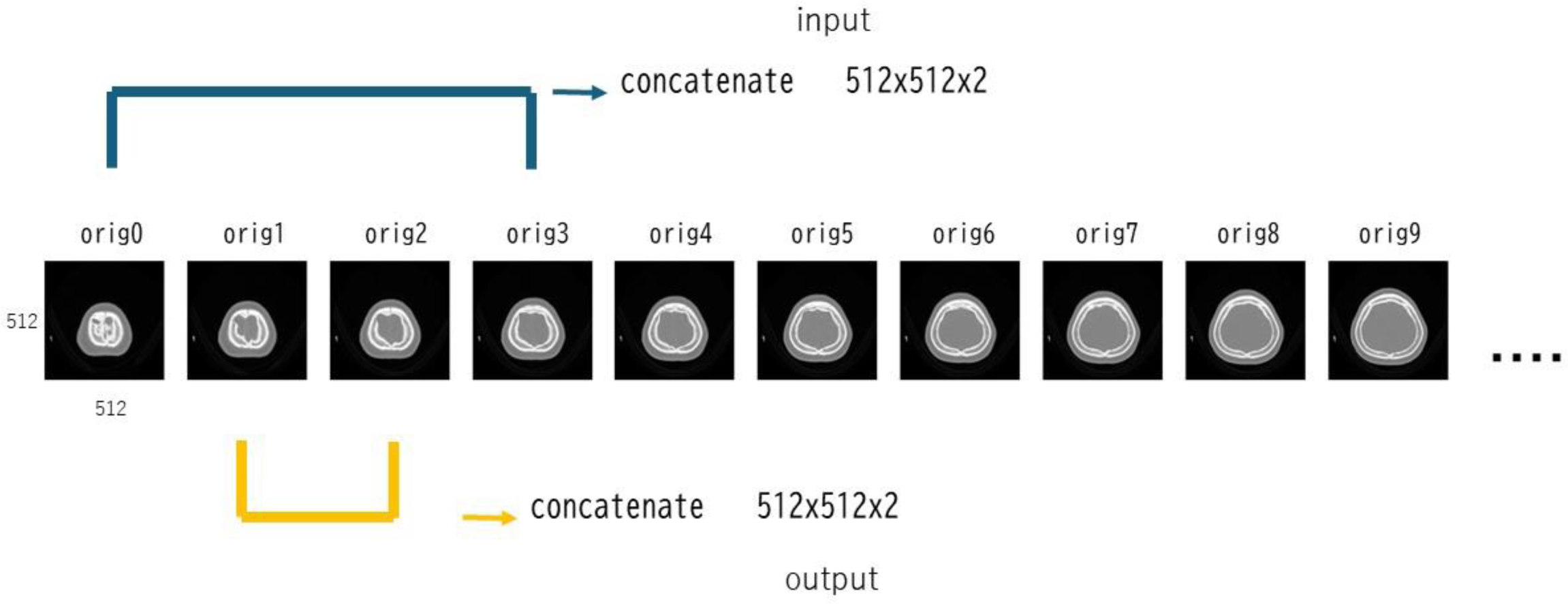
Making datasets for 1/3 re-slice system. An image and the fourth image, which skipped two images, were concatenated and stored for input data. The skipped two images were concatenated and stored for output data. Also, an image and the seventh are concatenated for input. The third and fifth were concatenated and stored for the corresponding output.

**Figure 3.**
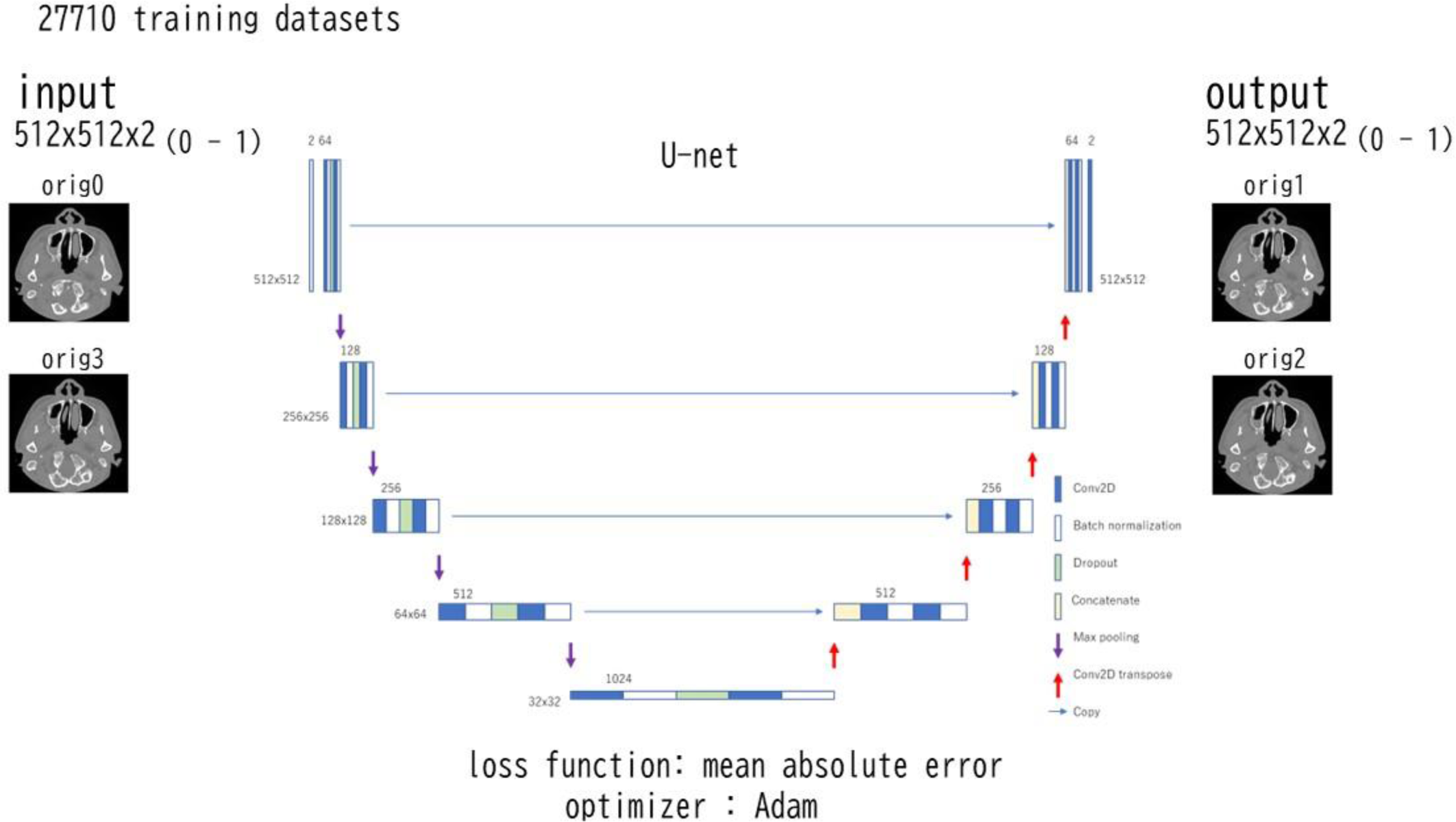
A U-net for 1/3 re-slice system. The input shape was 512x512x2 and the output shape was 512x512x2.

The 3rd and 5th images were stacked as output. The input/output dataset was divided into 70% (27710 pairs) for training and the rest (11877 pairs) for validation. A U-net(7) (https://pypi.org/project/keras-unet/) was customized as input: 512x512x2, output: 512x512x2. Data augmentation, (within rotation of 10 degrees, lateral and vertical shift of 10%, zoom of 20% and horizontal flip) was done with Keras ImageDataGenerator. Mean absolute error was utilized as the loss function. As the optimizer, Adam(8) was used. The U-net was trained with the training dataset for 200 epochs.

#### (2) **1/4 re-slice system** (Figure 4, 5)

One image and the 5th image (skipping 3 images) were paired and stacked (512x512x2). That was collected as input, and the 2nd, 3rd and 4th images stacked (512x512x3) were collected as output. Then, one image and the 9th image stacked were collected as input, the 3rd, 5th, and 7th images stacked as output. The input/output dataset was divided into 70% (27638 pairs) for training and the rest (11846 pairs) for validation. A U-net (input: 512x512x2, output: 512x512x3) was trained with the training dataset. Training method was the same as the 1/3re- slice system.

**Figure 4.**
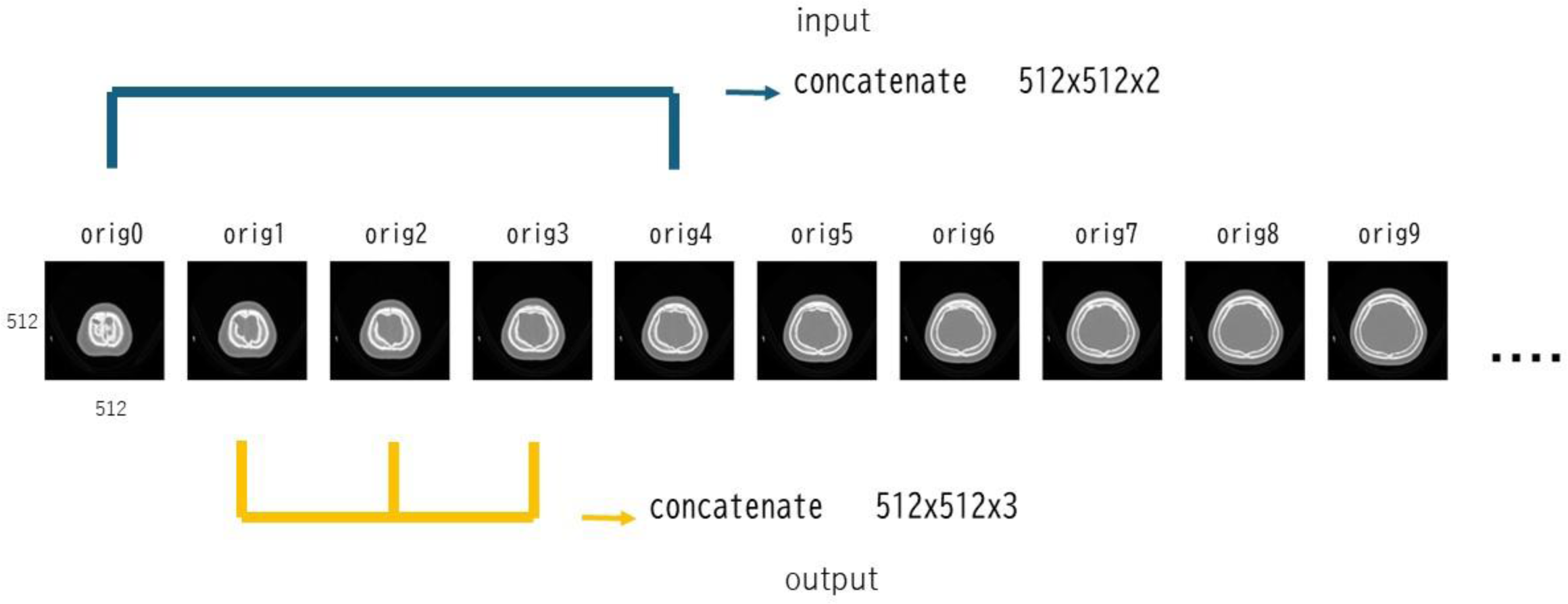
Making datasets for 1/4 re-slice system. An image and the fifth image, which skipped three images, were concatenated and stored for input data. The skipped three images were concatenated and stored for output data. An image and the 9th image were concatenated for input. The 3rd, 5th and 7th images were concatenated for the corresponding output.

**Figure 5.**
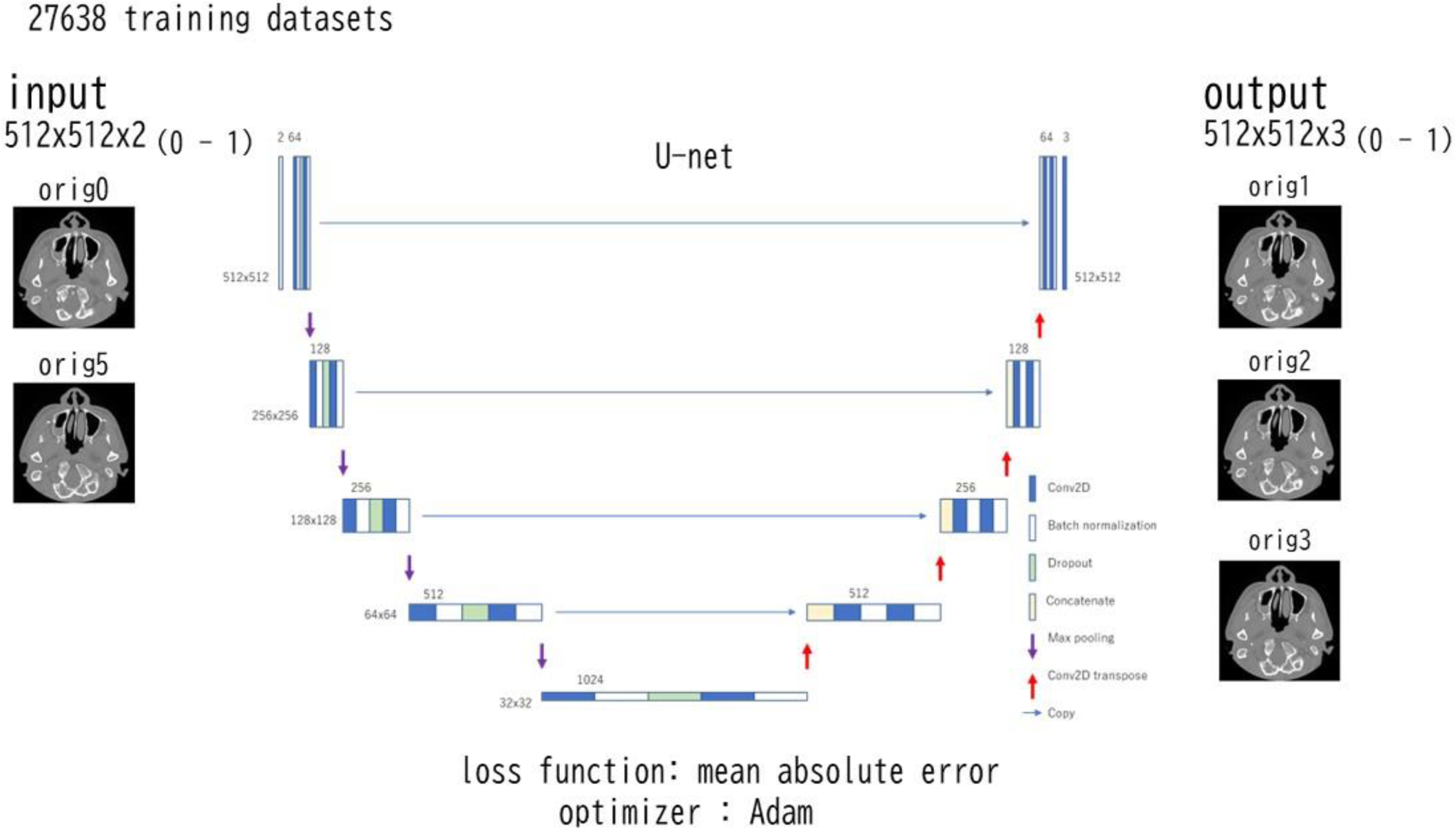
A U-net for 1/4 re-slice system. The input shape was 512x512x2 and the output shape was 512x512x3.

#### (3) 1/5 re-slice system (Figure 6, 7)

One image and the 6th image (skipping 4 images) were paired and stacked (512x512x2). It was collected as input, and the 2nd, 3rd, 4th and 5th images stacked (512x512x4) were collected as output. Then, one image and the 11th image stacked was collected as input, and the 3rd, 5th, 7th and 9th images stacked as output. The input/output dataset was divided into 70% (27496 pairs) for training and the rest (11785 pairs) for validation, and trained on a U-net (input: 512x512x2, output: 512x512x4). Training method was the same as the 1/3re-slice system.

**Figure 6.**
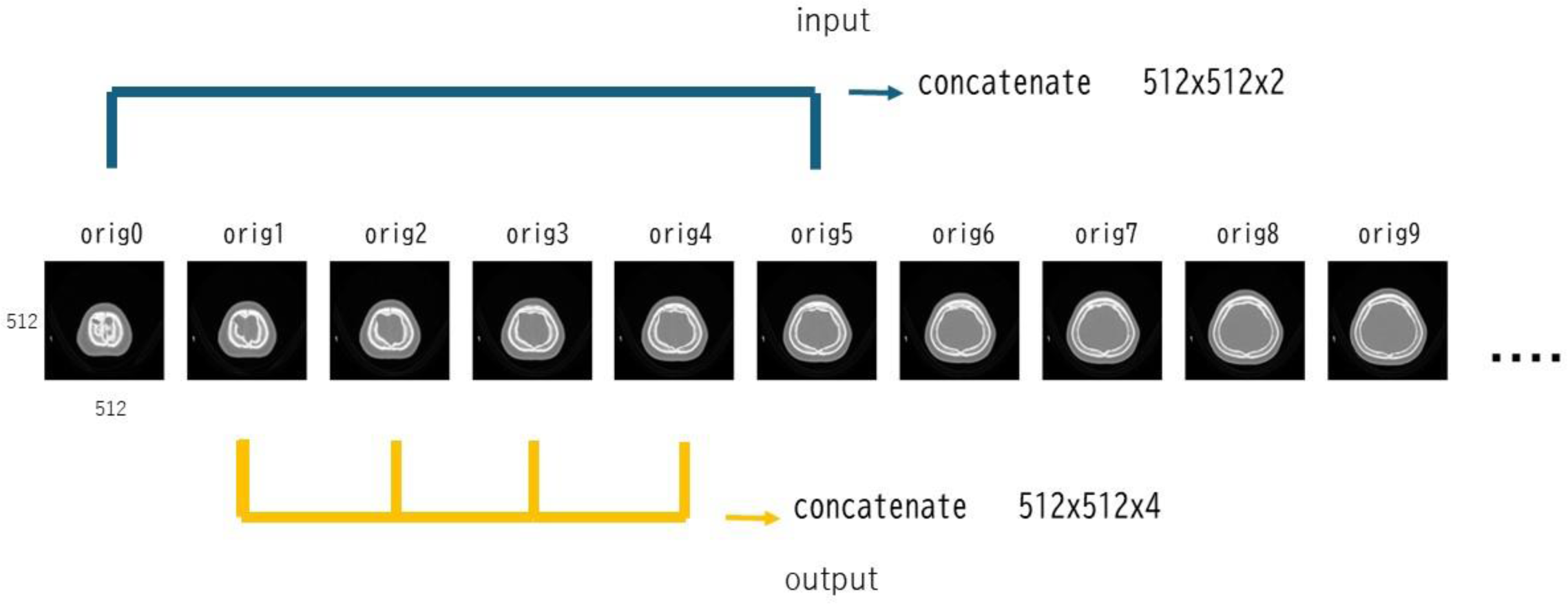
Making datasets for 1/5 re-slice system. An image and the sixth image, which skipped four images, were concatenated and stored for input data. The skipped four images were concatenated and stored for output data. An image and the 11th image were concatenated for input. The 3rd, 5th, 7th and 9th images were concatenated for the corresponding output.

**Figure 7.**
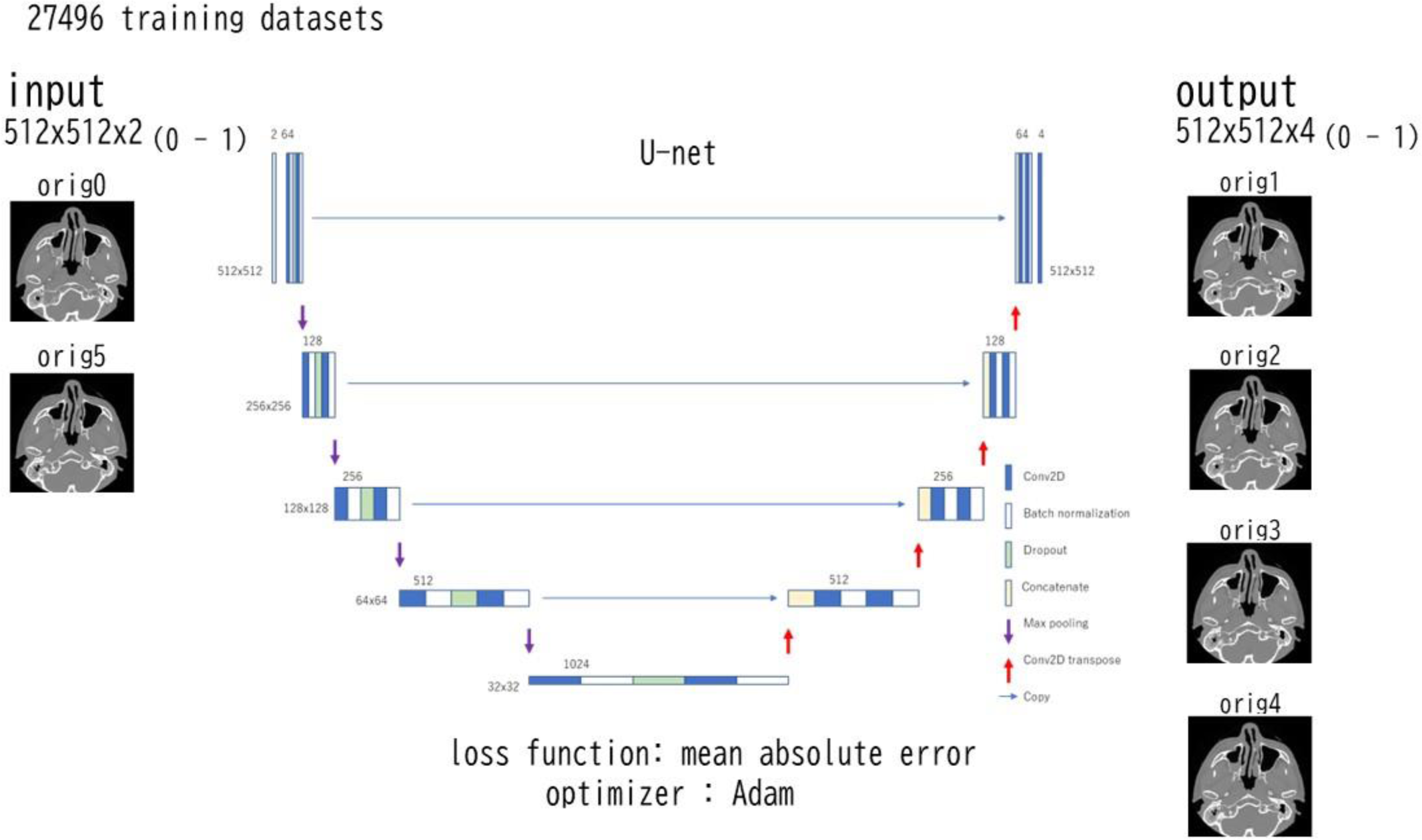
A U-net for 1/4 re-slice system. The input shape was 512x512x2 and the output shape was 512x512x4.

### Validation

#### Linear interpolation

With validation datasets (paired and stacked two images), linear interpolation images were made by calculating weighted average, in accordance with the distance from the original two images.

#### U-net prediction

Validation datasets were fed to the trained U-nets and the images were created.

#### Similarity index

With scikit-image library(9) (https://scikit-image.org/), peak signal to noise ratio (PSNR) between the created image and the corresponding original image was calculated with the data range of 1.0. Structural similarity (SSIM), with the data range of 1.0, and mean squared error (MSE) were also computed using the library. These computed similarity indexes were compared with paired-t analysis using SciPy library(10) (https://scipy.org/).

## Results

Comparison of similarity indexes (PSNR, SSIM, MSE) in linear interpolation and U-net prediction with the corresponding original image is summarized in Table 2. Paired-t analysis statistically revealed the significant difference between them in all indexes.

Examples of comparison between the images original, created by linear interpolation and U-net prediction are shown in Figure 8 – 13.

**Figure 8.**
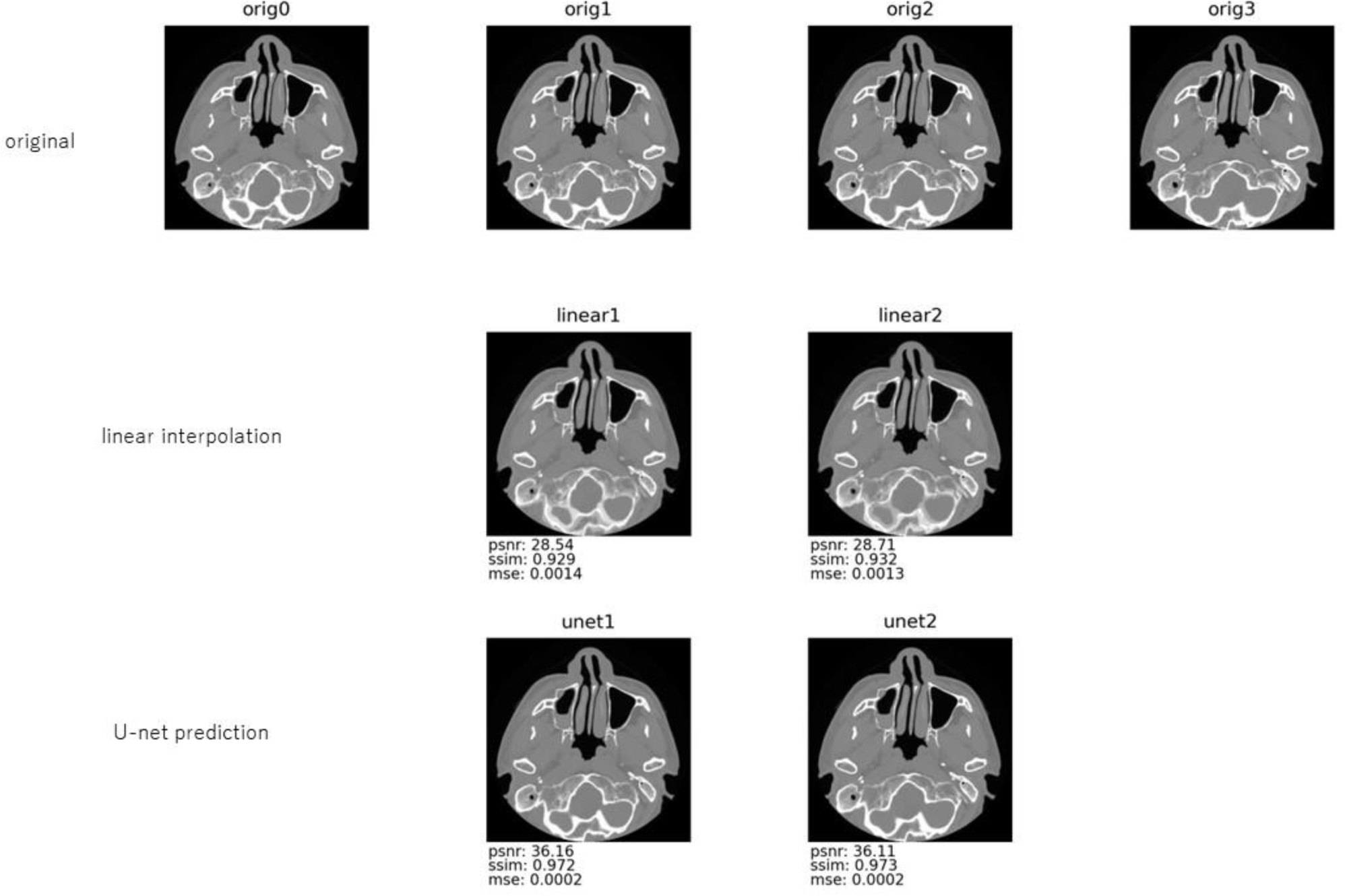
An example of comparison between the images original, created by linear interpolation and U-net prediction (1/3 re-slice system). The first row: A series of CT slices with an interval of 0.5mm. The interval between orig0 and orig3 was 1.5mm. The second row: Images created by linear interpolation of orig0 and orig3. The third row: Images created by U-net prediction with the input of orig0 and orig3. psnr: peak signal-to-noise ratio, ssim: structural similarity and mse: mean squared error were calculated in comparison with the corresponding image of the first row.

**Figure 9.**
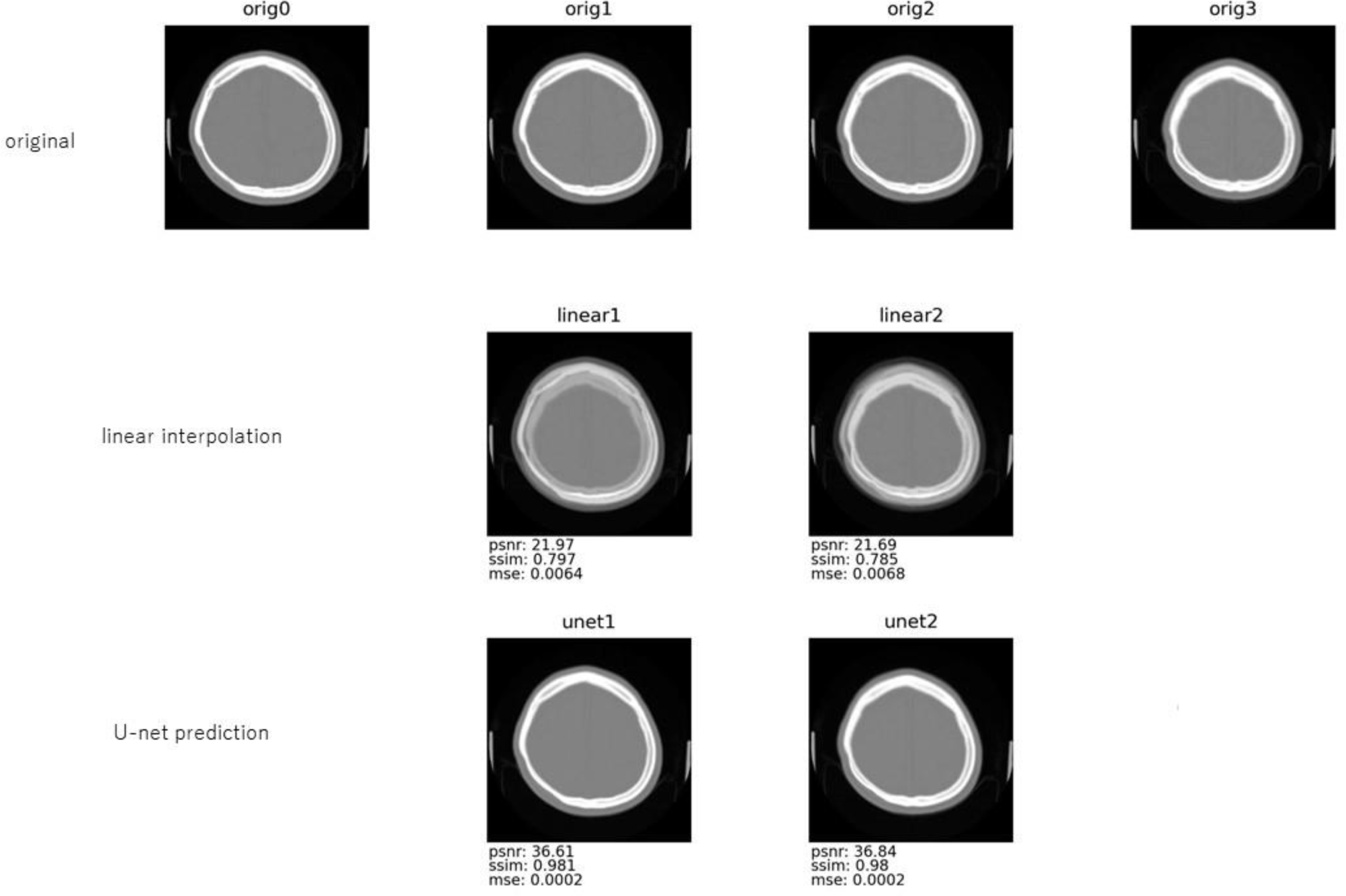
An example of comparison between the images original, created by linear interpolation and U-net prediction (1/3 re-slice system). The first row: A series of CT slices with an interval of 4mm. The interval between orig0 and orig3 was 12mm. The second row: Images created by linear interpolation of orig0 and orig3. The third row: Images created by U-net prediction with the input of orig0 and orig3. psnr: peak signal-to-noise ratio, ssim: structural similarity and mse: mean squared error were calculated in comparison with the corresponding image of the first row.

**Figure 10.**
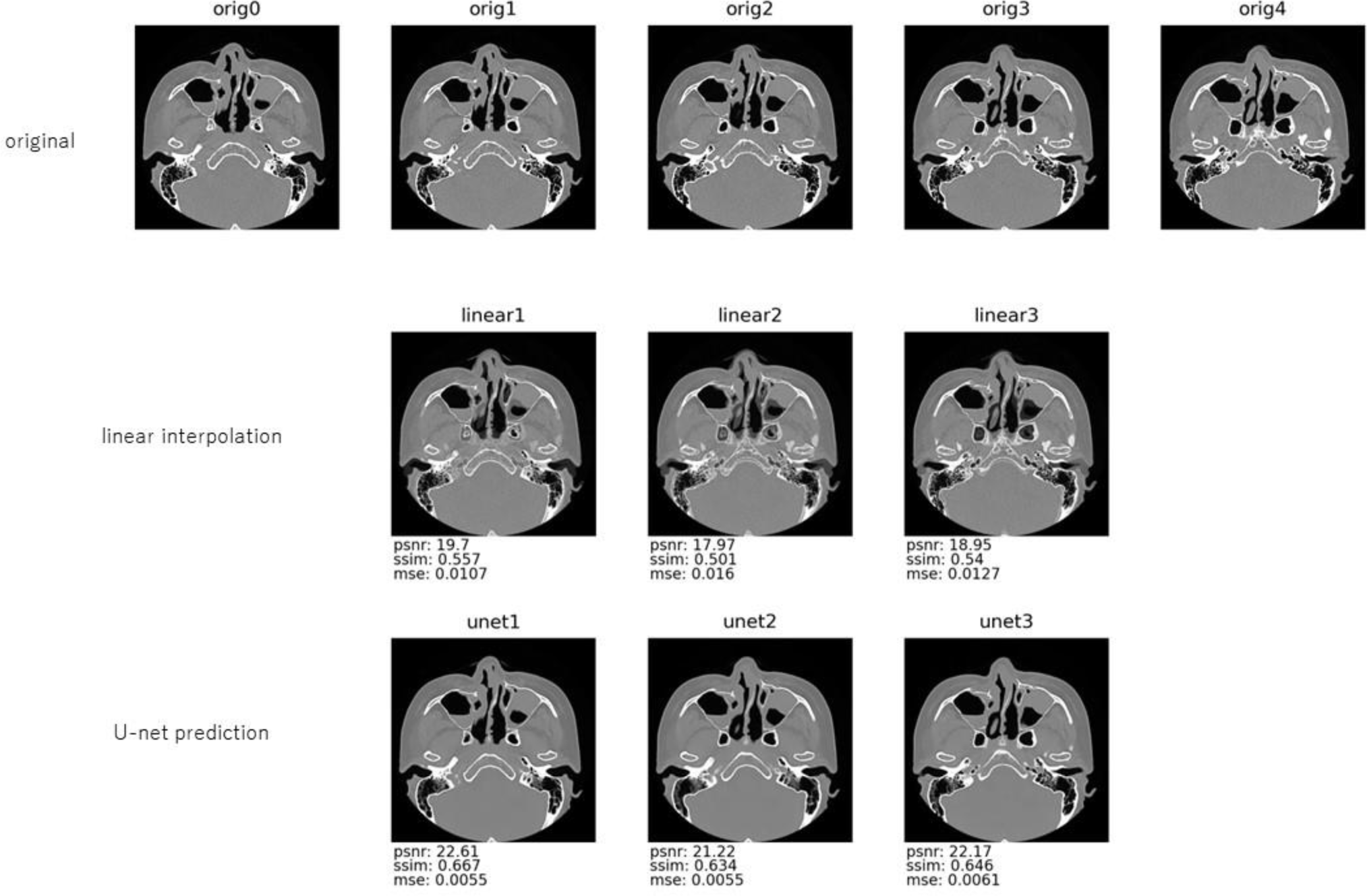
An example of comparison between the images original, created by linear interpolation and U-net prediction (1/4 re-slice system). The first row: A series of CT slices with an interval of 1mm. The interval between orig0 and orig4 was 4mm. The second row: Images created by linear interpolation of orig0 and orig4. The third row: Images created by U-net prediction with the input of orig0 and orig4. Computed similarity indexes were better in U-net prediction than in linear interpolation. psnr: peak signal-to-noise ratio, ssim: structural similarity and mse: mean squared error were calculated in comparison with the corresponding image of the first row.

**Figure 11.**
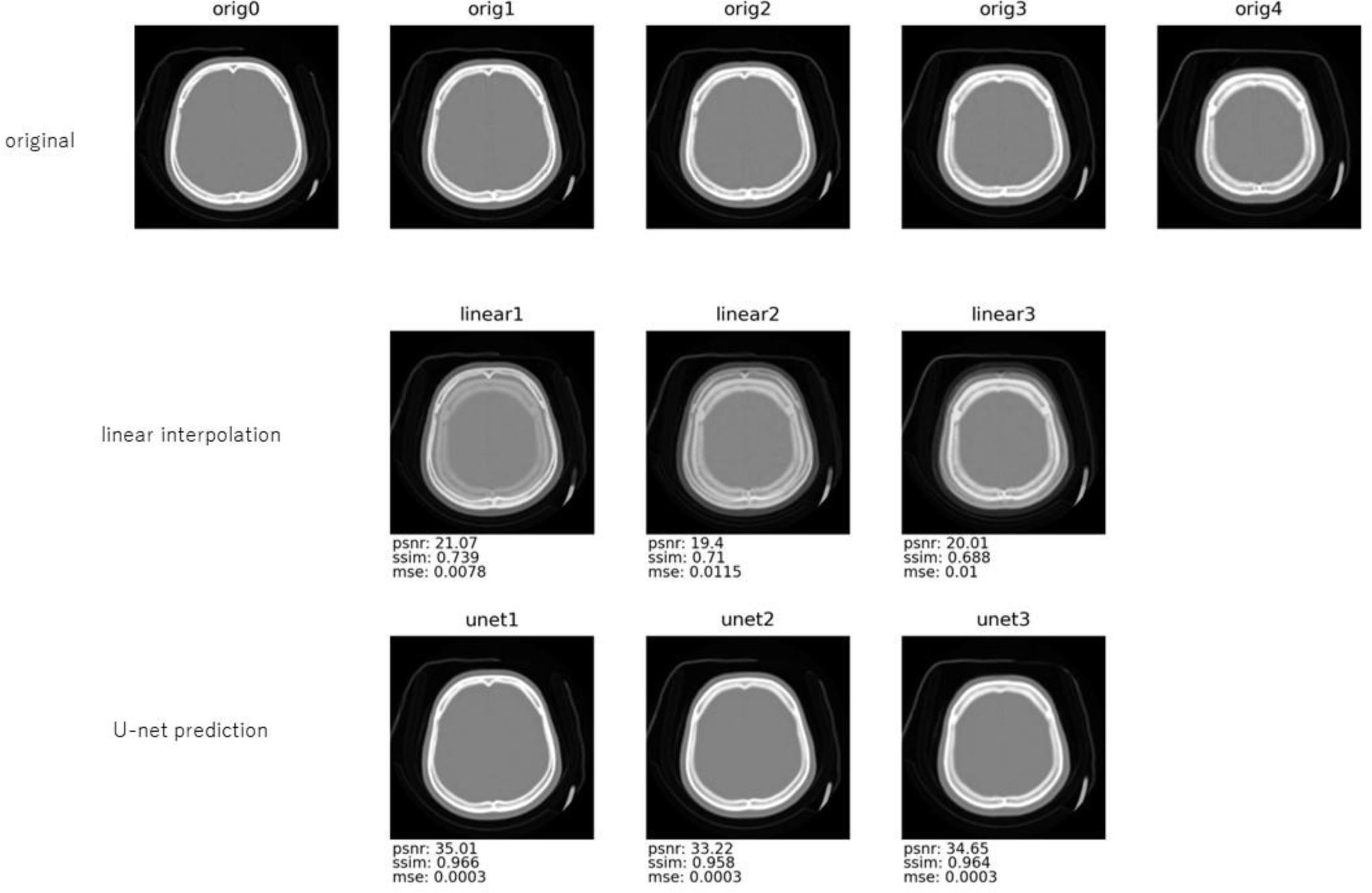
An example of comparison between the images original, created by linear interpolation and U-net prediction (1/4 re-slice system). The first row: A series of CT slices with an interval of 4mm. The interval between orig0 and orig4 was 16mm. The second row: Images created by linear interpolation of orig0 and orig4. The third row: Images created by U-net prediction with the input of orig0 and orig4. psnr: peak signal-to-noise ratio, ssim: structural similarity and mse: mean squared error were calculated in comparison with the corresponding image of the first row.

**Figure 12.**
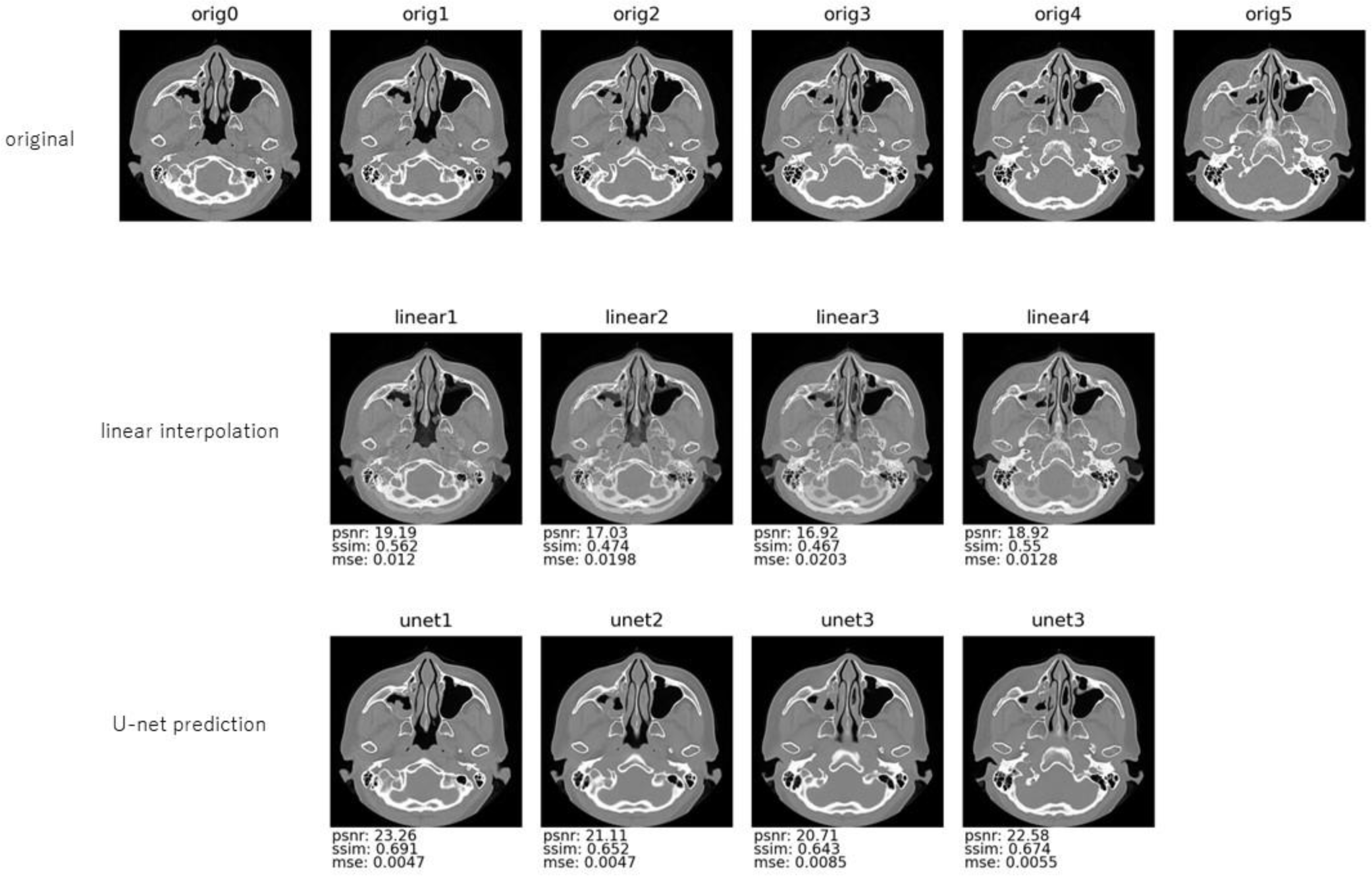
An example of comparison between the images original, created by linear interpolation and U-net prediction (1/5 re-slice system). The first row: A series of CT slices with an interval of 1mm. The interval between orig0 and orig5 was 5mm. The second row: Images created by linear interpolation of orig0 and orig5. The third row: Images created by U-net prediction with the input of orig0 and orig5. psnr: peak signal-to-noise ratio, ssim: structural similarity and mse: mean squared error were calculated in comparison with the corresponding image of the first row.

**Figure 13.**
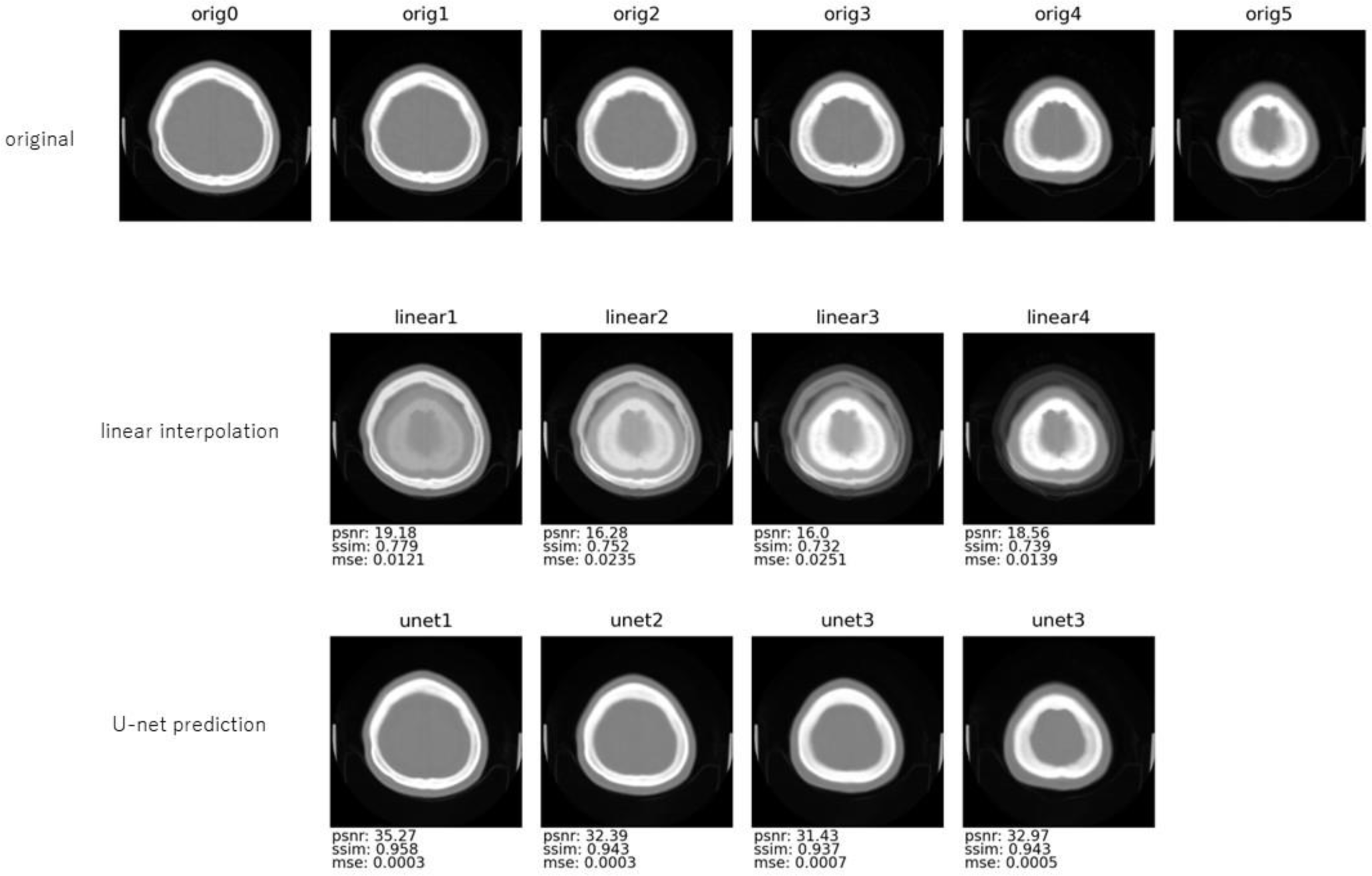
An example of comparison between the images original, created by linear interpolation and U-net prediction (1/5 re-slice system). The first row: A series of CT slices with an interval of 4mm. The interval between orig0 and orig5 was 20mm. The second row: Images created by linear interpolation of orig0 and orig5. The third row: Images created by U-net prediction with the input of orig0 and orig5. psnr: peak signal-to-noise ratio, ssim: structural similarity and mse: mean squared error were calculated in comparison with the corresponding image of the first row.

Fractured parts were well reproduced for human eye.

There were some datasets in which U-net prediction presented worse similarity indexes than linear interpolation. Two examples of them are shown in Figure 14, 15. Metal artifact can be seen in the original images of them.

**Figure 14.**
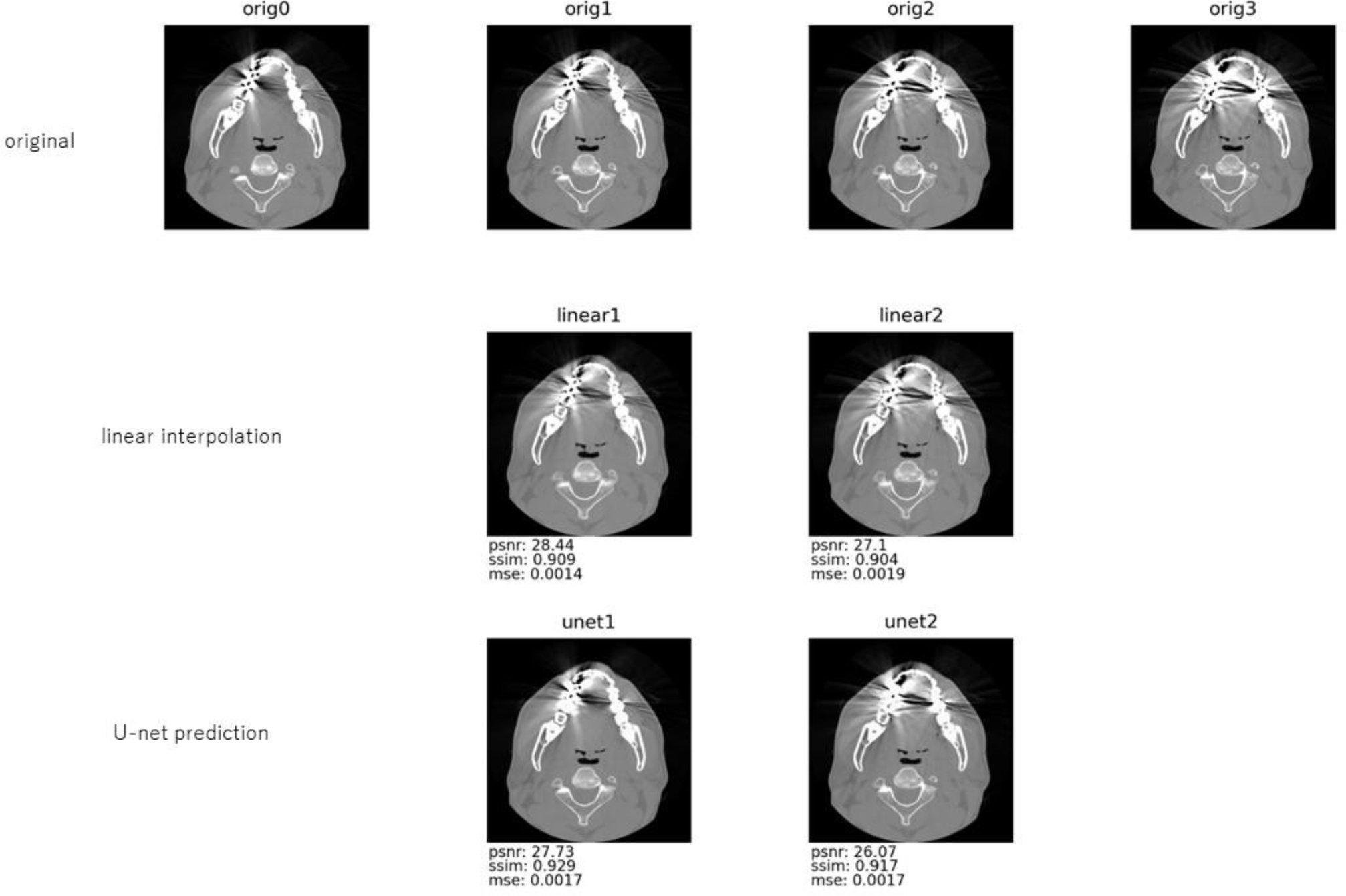
An example of dataset, U-net prediction showed worse peak signal-to-noise ratio and mean squared error than linear interpolation (1/3 re-slice system). Metal artifact was seen in the original images. The first row: A series of CT slices with an interval of 0.5mm. The interval between orig0 and orig3 was 1.5mm. The second row: Images created by linear interpolation of orig0 and orig3. The third row: Images created by U-net prediction with the input of orig0 and orig3. psnr: peak signal-to-noise ratio, ssim: structural similarity and mse: mean squared error were calculated in comparison with the corresponding image of the first row.

**Figure 15.**
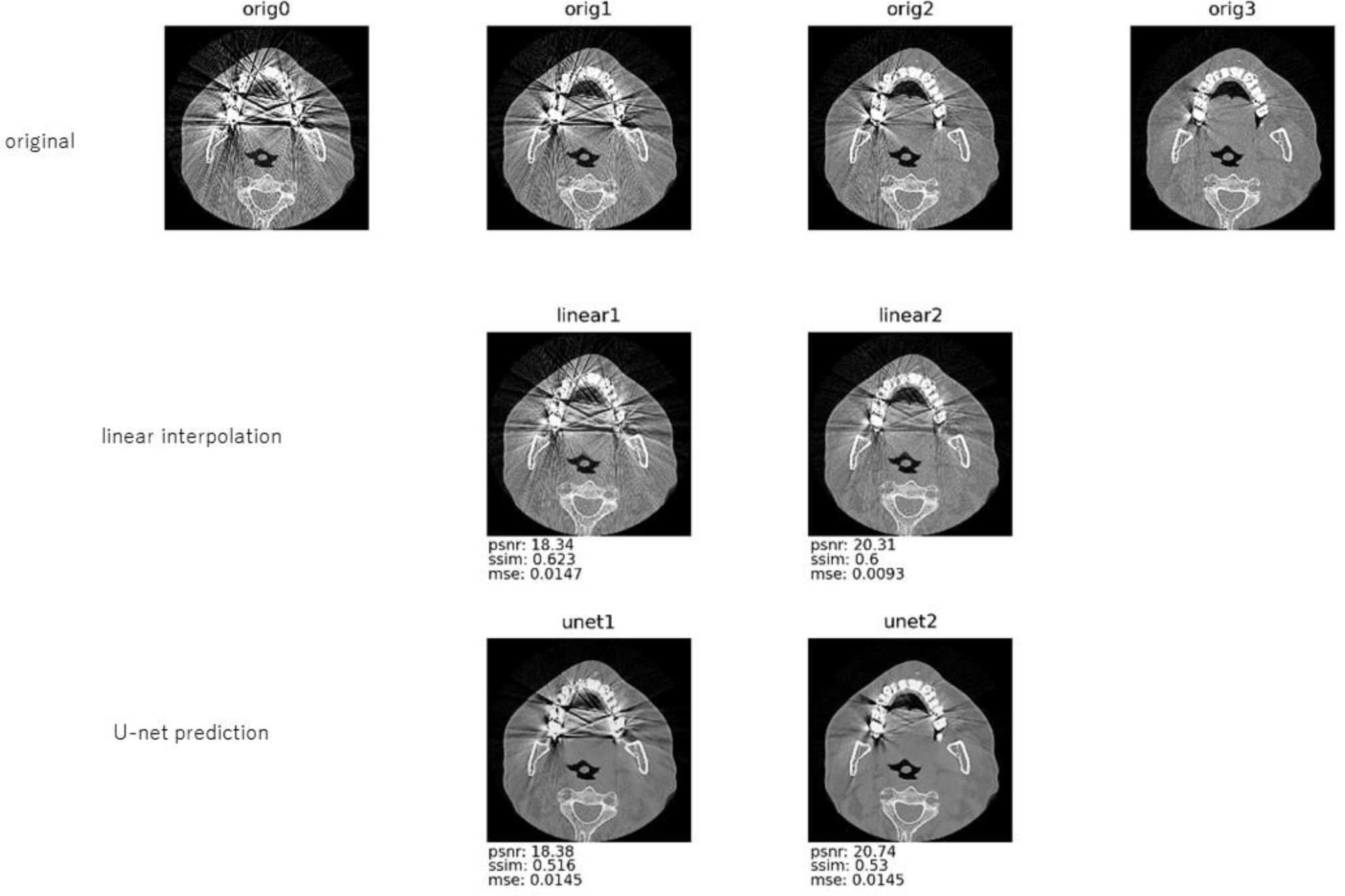
An example of dataset, U-net prediction showed worse structural similarity than linear interpolation (1/3 re-slice system). Metal artifact was seen in the original images. The first row: A series of CT slices with an interval of 0.5mm. The interval between orig0 and orig3 was 1.5mm. The second row: Images created by linear interpolation of orig0 and orig3. The third row: Images created by U-net prediction with the input of orig0 and orig3. psnr: peak signal-to-noise ratio, ssim: structural similarity and mse: mean squared error were calculated in comparison with the corresponding image of the first row.

## Discussion

Linear interpolation is a simple method to create an intermediate image from two given images. Calculation of the linear weighted average of pixel value in the same position of the images are done. The method can be performed quickly because of its simplicity. The transition is done without spatial warping, so the intermediate images are the simple mixtures of the original two images. Spline interpolation can be another option. At least three points are required to define a spline curve. Polynomials are used to determine the pixel values from the corresponding pixels. Calculation needs time and physical resources.

Image morphing can be defined as the process of constructing image sequence that illustrate gradual transition between the two images(11)(12). This technique involves warping, changing the position of key points and deforming the images. The transition gives an observer realistic and spectacular impression. In the entertainment industry, image morphing is used very often. Establishing correspondence of two images with pairs of feature points is the beginning of the process. Although many attempts have been done to do this procedure automatically, manual selection is usually adopted.

There was a report using an image morphing technique to create images between CT and NMR scans(13). They automatically extracted features partly. They could not fully automate the process of corresponding points between the images. And it is assumed that using morphing technique pair by pair takes time.

Kudo et al. utilized three dimensional conditional generative adversarial networks to produce super-resolution CT images(14). Jurek et al. constructed a convolutional neural network to reconstruct super-resolution MR images(15). As three-dimensional networks demand large computation resource, former cropped 160 x 160 x 160 and the latter 180 x180 x 180 pixel region.

In this study, three U-nets were trained individually for 1/3, 1/4 and 1/5 re-slicing system. We considered that these 3 systems would be sufficient for our practical use. For example, to obtain 2mm interval series of CT images from 4mm slice width, it can be done by taking one out every other from the products by the 1/4 re-slice system. To obtain 1mm interval series of CT images from 0.625mm slice width, it can be done by taking out every 8th from the products by the 1/5 re- slice system, which makes 0.125mm slice width.

The images generated by the trained U-nets showed statistically better similarity indexes to the original ones than linear interpolation. Though, with the datasets including metal artifact, some reversal occurred. The frequency of it was low.

How similar the product should be is hard discuss. It depends on the quality of demand, cost, physical resource, time and so on. This study was to make intermediate slice images from the existing two images.

The datasets used included variation of slice width as shown in Table 1. The datasets with less variation may have achieved better results. Though, that may have reduced robustness. We would like to evaluate our approach practical.

**Table 1.** Variation of the slice thickness in the CT image series.

| slice thickness (mm) | number |
| --- | --- |
| 0.5 | 25 |
| 0.625 | 11 |
| 1 | 8 |
| 2 | 35 |
| 2.5 | 1 |
| 3 | 4 |
| 4 | 4 |
| 5 | 14 |
| total | 102 |

**Table 2.**
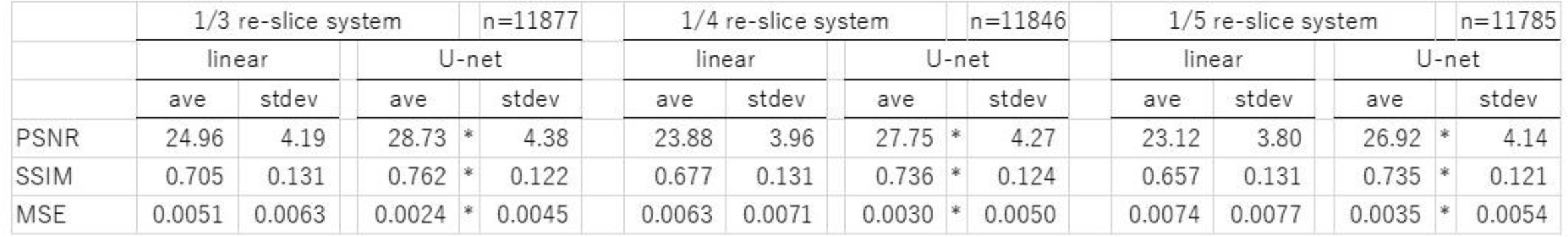
Comparison of image similarity indexes. PSNR: peak signal-to-noise ratio, SSIM: structural similarity and MSE: mean squared error were calculated in comparison with the corresponding original image. * p<0.001 by paired-t analysis

It cannot be denied that a series of CT images, obtained with thin slice thickness, provide more detailed information than that with thicker ones. Sometimes, it is discussed that thinly re-sliced images virtually from thicker sparse images can overcome this shortcoming(14). It is true that virtual re-slicing will increase image resolution vertically or three dimensionally. They appear clearer and more detailed to human eyes. Though, it does not mean that the system let real detailed information visible which was originally invisible in thicker slice. The information, not contained in the original data, may not be the real. If new information appears, no matter what method is used, it cannot be denied that it is false. This point should not be forgotten. Nakamoto et al. reported their experience in using virtual thin slice system as practitioners(16). The diagnostic ability for vertebral compression fracture was impaired by the system.

## Conclusion

Three re-slice systems utilizing U-nets were developed. Comparing with linear interpolation method, the systems were able to generate statistically better inter-slice images from two existing CT images.

## Data Availability

All data produced in the present study are available upon reasonable request to the authors

## Acknowledgement

Authors thank members who cooperated in accumulation of the CT images for Japanese Facial Bone Fracture CT Collection Project: Yuki Suzuki, Shoji Kido, Takeo Miki, Nobuyuki Mitsukawa, So Moriyama, Michiko Fukuba, Yuzo Komuro, Naoya Oshima, Shunsuke Yuzuriha, Yusuke Shikano, Tateki Kubo, Shotaro Suzuki, Satoshi Takagi, Haruna Yoshiike.

There is no financial conflict of interest.

